# Conditionality of COVID-19 vaccine acceptance in European countries

**DOI:** 10.1101/2021.06.24.21259453

**Authors:** Leonardo W Heyerdahl, Muriel Vray, Benedetta Lana, Nastassia Tvardik, Nina Gobat, Marta Wanat, Sarah Tonkin-Crine, Sibyl Anthierens, Herman Goossens, Tamara Giles-Vernick

## Abstract

The COVID-19 vaccine rollout in recent months offers a powerful preventive measure that may help control SARS-CoV-2 transmission. Nevertheless, long-standing public hesitation around vaccines has heightened public health concerns that vaccine coverage may not achieve desired public health impacts.This cross-sectional survey was conducted online in December 2020 among 7000 respondents (aged 18 to 65) in Belgium, France, Germany, Italy, Spain, Sweden, and Ukraine. The survey included open text boxes for fuller explanation of responses. Projected COVID-19 vaccine coverage varied and may not be sufficiently high among certain populations to achieve herd immunity. Overall, 56.9% would accept a COVID-19 vaccine, 19.0% would not, and 24.1% did not know or preferred not to say. By country, between 44% (France) and 66% (Italy) of respondents would accept a COVID-19 vaccine. Respondents expressed conditionality in open responses, voicing concerns about vaccine safety and mistrust of authorities. Public health campaigns must tackle these safety concerns.

**Highlights:**

- Mixed-method survey studied expected COVID vaccine uptake in 7 European countries.
- Projected COVID vaccine acceptance by country ranged from 44% to 66%.
- Explicit COVID vaccine acceptance or rejection was conditional.
- Study finds concerns about vaccine safety and authorities’ competence and honesty.
- Vaccine communications should address safety anxieties and target specific groups.

---

The COVID-19 vaccine rollout offers one powerful measure that can contribute to pandemic control. A successful rollout, however, requires a coordinated approach enhancing trust and acceptance of these vaccines, as well as equitable access, particularly for those groups who most need protection[1]. Several studies in Europe and beyond have evaluated whether members of the public would accept a COVID-19 vaccine; projected vaccine uptake figures have heightened concerns that vaccine coverage may fall short of desired public health impacts [2-6].

Vaccine acceptance is a complex, “multi-layered” process [7], influenced by contextual factors [8, 9] that include past vaccine experience, shared perceptions of disease severity[10], experiences with the health system[11], and trust in authorities[12]. The newly-developed, approved COVID-19 vaccines have made the decision process even more complex, despite health authorities’ efforts to reassure European publics about vaccine efficacy and safety[2]. These concerns are sufficiently complex and dynamic that they cannot easily be distilled into closed questions about projected vaccine acceptance and related factors.

Just prior to vaccine rollout in Europe, we conducted a mixed-method online study[13], whose primary objective was to estimate self-reported COVID-19 vaccine acceptance in seven European countries, and to identify factors associated with vaccine hesitancy.

## Methods

This cross-sectional survey, conducted by the market research firm Ipsos, sought to estimate expected COVID-19 vaccine acceptance and to evaluate factors associated with vaccine hesitancy.

The study was implemented through an online survey from December 4 to 16, 2020 among 7000 respondents in Belgium, France, Germany, Italy, Spain, Sweden, and Ukraine. Ipsos employed 1000-person panels in each country, between the ages of 18 and 65, imposing quotas by age, gender, geographical region, and working status, aligned with nationally-representative proportions of this age range.

The following quantitative data were collected among respondents: socio-economic and demographic characteristics; projected COVID-19 vaccine acceptance; trust in sources of medical and scientific information; trust in national, European, and international institutions and authorities, as well as in pharmaceutical companies; perception of vaccine contents, purposes, and safety; and political affiliation.

The survey also collected open text responses from respondents to elaborate results from the core quantitative study, namely whether they would accept COVID-19 vaccination.

All qualitative variables were expressed as percentage. Responses to acceptance of vaccine (yes/no/don’t know) were compared using χ2 test for qualitative variables. A p-value ≤0.05 was considered statistically significant. Data were analyzed using STATA Software Version 15.1 (Stata Corporation, College Station, Texas, USA).

NVivo software (Windows Release 1, QSR International) supported the qualitative data analysis. We evaluated 2251 text responses from the French, Italian, and Spanish panels, because vaccine hesitancy and refusal have been prominent in these countries [2] and conducted deductive and inductive coding to develop a thematic analysis of responses.

The University of Antwerp ethics committee provide ethical approval (20/13/150). All participants furnished informed consent before participating in the survey.

## Results

Table 1 summarizes the social and demographic profiles of survey respondents. Although respondent panels for each country were representative in terms of age (18-65 years), gender, occupational status, and country region, the panels were heavily weighted towards those with higher than primary-level education.

**Table 1.** Characteristics of study population.

|  | **Total**<br>**N (%)** | **Belgium**<br>**N (%)** | **France**<br>**N (%)** | **Germany**<br>**N (%)** | **Italy**<br>**N (%)** |
| --- | --- | --- | --- | --- | --- |
| Women | 3 516 (50.3) | 498 (49.8) | 511 (51.2) | 497 (49.9) | 504 (50.4) |
| Age |  |  |  |  |  |
| 18-24 | 916 (13.1) | 132 (13.2) | 134 (13.4) | 114 (11.4) | 110 (11.0) |
| 25-34 | 1 455 (20.8) | 206 (20.6) | 200 (20.0) | 192 (19.2) | 187 (18.7) |
| 35-44 | 1 546 (22.1) | 215 (21.5) | 218 (21.8) | 209 (20.9) | 248 (24.8) |
| 45-54 | 1 640 (23.4) | 233 (23.3) | 224 (22.4) | 259 (25.9) | 243 (24.3) |
| 55-65 | 1 443 (20.6) | 214 (21.4) | 224 (22.4) | 226 (22.6) | 212 (21.2) |
| Marital status |  |  |  |  |  |
| Single | 2 866 (40.9) | 448 (44.8) | 373 (37.3) | 488 (48.8) | 426 (42.6) |
| Married/<br>Domestic<br>Partner | 4 134 (59.1) | 552 (55.2) | 627 (62.7) | 512 (51.2) | 574 (57.4) |
| Employment |  |  |  |  |  |
| Working* | 4 548 (65.0) | 641 (64.1) | 661 (66.1) | 746 (74.6) | 568 (56.8) |
| Not working | 2 452 (35.0) | 359 (35.9) | 339 (33.9) | 254 (25.4) | 435 (43.5) |
| Education |  |  |  |  |  |
| Primary | 479 (6.8) | 126 (12.6) | 5 (0.5) | 37 (3.7) | 81 (8.1) |
| Secondary | 3 234 (46.2) | 350 (35.0) | 412 (41.2) | 612 (61.2) | 669 (66.9) |
| Tertiary | 3 287 (47.0) | 524 (52.4) | 583 (58.3) | 351 (35.1) | 250 (25.0) |
|  | **Spain**<br>**N (%)** | **Sweden**<br>**N (%)** | **Ukraine**<br>**N (%)** |  |  |
| Women | 498 (49.8) | 507 (50.7) | 501 (50.1) |  |  |
| Age |  |  |  |  |  |
| 18-24 | 108 (10.8) | 157 (15.7) | 161 (16.1) |  |  |
| 25-34 | 212 (21.2) | 214 (21.4) | 244 (24.4) |  |  |
| 35-44 | 261 (26.1) | 180 (18.0) | 215 (21.5) |  |  |
| 45-54 | 229 (22.9) | 225 (22.5) | 227 (22.7) |  |  |
| 55-65 | 190 (19.0) | 224 (22.4) | 153 (15.3) |  |  |
| Marital status |  |  |  |  |  |
| Single | 394 (39.4) | 399 (39.9) | 338 (33.8) |  |  |
| Married/<br>Domestic<br>Partner | 606 (60.6) | 601 (60.1) | 662 (66.2) |  |  |
| Employment |  |  |  |  |  |
| Working* | 565 (56.5) | 756 (75.6) | 611 (61.1) |  |  |
| Not working | 435 (43.5) | 244 (24.4) | 389 (38.9) |  |  |
| Education |  |  |  |  |  |
| Primary | 106 (10.6) | 72 (7.2) | 52 (5.2) |  |  |
| Secondary | 238 (23.8) | 549 (54.9) | 404 (40.4) |  |  |
| Tertiary | 656 (65.6) | 379 (37.9) | 544 (54.4) |  |  |

\*Defined as full-time employment, part-time employment or self-employment

Figure 1 shows the numbers and percentages of those who would accept or reject vaccination, or who do not know, globally and by country. Overall, 3983 (56.9%) would accept vaccination, 1325 (19.0%) would not, and 1688 (24.1%) did not know or preferred not to say. Survey results show that projected COVID-19 vaccine acceptance varied across countries, and in some populations would be insufficient to achieve herd immunity (Table 2). Between 44% (n=441, France) and 66% (n=1658, Italy) of respondents would **accept** a COVID-19 vaccine if it was safe, effective and provided free-of-charge. Among the respondents, however, between 21% (n=211, Italy) and 28% (n=279, France) did not know or preferred not to say if they would accept a vaccine.

**Table 2.** Factors linked to accepting or rejecting COVID-19 vaccination: n (%)

|  | Yes<br>(N=3 985) | No<br>(N=1 327) | Don't know/<br>Prefer not to say<br>(N=1 688) | Total<br>(N=7 000) | p-value |
| --- | --- | --- | --- | --- | --- |
| **COVID-19 vaccine** |  |  |  |  |  |
| Vaccine not |  |  |  |  |  |
| Dangerous<sup>1</sup> | 2 140 (53.7) | 216 (16.3) | 323 (19.1) | 2 679 (38.3) | p<0.001 |
| Safety considerations |  |  |  |  |  |
| Bypassed<sup>2</sup> | 1 408 (35.3) | 723 (54.5) | 766 (45.4) | 2 897 (41.4) | p<0.001 |
| Adjuvants unsafe<sup>3</sup> | 670 (16.8) | 568 (42.8) | 402 (23.8) | 1 640 (23.4) | p<0.001 |
| Microchips in vaccine<sup>4</sup> |  |  |  |  |  |
| Agree | 452 (11.3) | 338 (25.5) | 191 (11.3) | 981 (14.0) | p<0.001 |
| **Trust in** |  |  |  |  |  |
| National government<sup>5</sup> | 1 823 (45.8) | 239 (18.0) | 379 (22.5) | 2 441 (34.9) | p<0.001 |
| Pharmaceutical cos.<sup>6</sup> | 1 868 (46.9) | 232 (17.5) | 386 (22.9) | 2 486 (35.5) | p<0.001 |
| Physicians<sup>7</sup> | 3 414 (85.7) | 830 (62.6) | 1 274 (75.5) | 5 518 (78.8) | p<0.001 |
| Nurses<sup>7</sup> | 2 992 (75.1) | 768 (57.9) | 1 146 (67.9) | 4 906 (70.1) | p<0.001 |
| Pharmacists<sup>7</sup> | 2 837 (71.2) | 652 (49.1) | 1 010 (59.8) | 4 499 (64.3) | p<0.001 |
| Media (general)<sup>7</sup> | 2 093 (52.5) | 385 (29.0) | 613 (36.3) | 3 091 (44.2) | p<0.001 |
| Friends and Family<sup>7</sup> | 1 024 (25.7) | 344 (25.9) | 379 (22.5) | 1 747 (25.0) | p<0.024 |
| Social networks<sup>7</sup> | 536 (13.5) | 175 (13.2) | 187 (11.1) | 898 (12.8) | p<0.046 |
| **Political affiliation<sup>8</sup>** |  |  |  |  |  |
| Left | 642 (66.3) | 146 (15.1) | 181 (18.7) | 969 (13.8) |  |
| Center | 1 814 (60.0) | 527 (17.4) | 684 (22.6) | 3025 (43.2) | p<0.001 |
| Right | 936 (59.0) | 341 (21.5) | 310 (19.5) | 1587 (22.7) |  |
| NA* | 593 (41.8) | 313 (22.1) | 513 (36.2) | 1419 (20.3) |  |

<sup>1</sup>Respondents were asked to what extent they agreed or disagreed with the following claim: “A COVID-19 vaccine will not be dangerous to human health.”
<sup>2</sup>Respondents were asked to what extent they agreed or disagreed with the following claim: “I believe that safety considerations are being bypassed in the development of COVID-19 vaccinations.”
<sup>3</sup>Respondents were asked to what extent they agreed or disagreed with the following: “Adjuvants (ingredients which cause more antibodies to be produced), contained in most vaccines, have negative effects on human health.”
<sup>4</sup>Respondents were asked to what extent they agreed or disagreed with the following: “Authorities want to insert microchips in the COVID-19 vaccine to impose control over people.”
<sup>5</sup>Respondents were asked to what extent they agreed or disagreed with the following: “The national government is **being honest** with its citizens when managing COVID-19 pandemic.”
<sup>6</sup>Respondents were asked to what extent they agreed or disagreed with the following: “Pharmaceutical companies that are doing research on COVID-19 would be **honest** about what they discover.”
<sup>7</sup>The following 6 categories trust in specific sources of information. Respondents were specifically asked to evaluate “To what extent do you consider the following to be trustworthy, or not, as sources of information about **scientific studies** concerning the origins, treatment, prevention, or consequences of COVID-19?”
<sup>8</sup>Respondents were asked to place their political beliefs on a 10-point scale from left to right.
\*NA = Missing data

Figure 1 shows responses by respondents’ socio-demographic characteristics. Overall, and in all countries except Italy, women expressed less intention and more uncertainty about accepting a COVID-19 vaccine than men (n=1820, 51.8%, versus n=2163, 62.1%, and n=986, 28% versus n=700, 20.1% not knowing/preferring not to say). The oldest age cohorts indicated that they were more likely to accept vaccination (45-54 y, n= 952, 58.1%, 55-65 y, n=907, 62.9%), except for in Italy and Sweden. Married respondents were more likely to accept vaccination (n=2422, 58.6% versus 1562, 54.5%), except in France and Italy; respondents with higher educational levels (n=1997, 60.8%) (except in Spain) and those working (n=2629, 57.8%) (except in Spain) were more likely to accept vaccination.

Analysis of factors associated with vaccine hesitancy focused on the safety and purported contents of the vaccine itself (Table 2). The safety of COVID-19 vaccines and their purported contents were significant factors correlating with projected acceptance, refusal, and not knowing. Across the seven countries, among the 3985 respondents who would accept the vaccine, 53.7% (n=2140) believed that COVID vaccines “would not be dangerous”, compared to 216 (16.3%) among the 1327 respondents who would not accept vaccination. That said, among respondents who would reject a COVID-19 vaccine, 54.5% believed that safety considerations had been bypassed in vaccine development, and 42.8% claimed that adjuvants were “dangerous to human health”, compared to 35.3% and 16.8%, respectively, among those who would accept vaccination. Among respondents who would not accept vaccination, 25.5% believed that authorities wanted to insert “microchips” in COVID-19 vaccinations to control European populations; yet up to 11.3% of respondents believing in this objective would nevertheless accept a COVID-19 vaccine.

Intentions to accept a COVID-19 vaccine were strongly correlated with respondents’ trust in their national governments (45.8% accepting vaccination, versus 18.0% rejecting and 22.5% not knowing or saying) and in pharmaceutical companies (46.9% accepting vaccination, versus 17.5% rejecting and 22.9% not knowing or saying). In addition, projected vaccine acceptance was also strongly linked to trust in physicians (85.7% (n=3414) accepting, 62.6% (n=830) rejecting, and 75.5% (n=1274) not knowing or saying) nurses (75.1% (n=2992) accepting, 57.9% (n=768) rejecting, 67.9% (n=1146) not knowing or saying) and pharmacists (71.2% (n=2837) accepting, 49.1% (n=652) rejecting, and 59.8% (n=1010) not knowing or saying) as sources of medical information. Respondents who identified their political affiliation as “left” were more likely to accept vaccination than those who identified with the “right” or those not responding to the question (66.3% versus 59.0% and 41.8%, respectively).

Qualitative data collected through this survey revealed further insight into why respondents would accept or reject a vaccine. Those who would accept a vaccine indicated that it would confer individual, familial and societal protection and restore daily life and economic activity. One respondent explained that vaccination was essential, noting, “I think this disease is terrible. It has taken away our freedom to live as we did before. We need to regain our freedom and our joy of living.” Those maintaining that they would refuse vaccination claimed that it was unnecessary: they did not believe they were at risk for COVID-19, took other health precautions, or thought that viral mutations would render a vaccine useless and unnecessary. “COVID is a virus that mutates all the time, observed another respondent, “which will probably render any vaccine that I receive useless.”

Most important, our qualitative analysis shows that projected COVID-19 vaccine acceptance and refusal were conditional. Respondents intending to accept vaccination claimed that they would only do so if it was proven to be safe, which took considerable time to demonstrate. “If a real scientific studied indicated that it was effective and certain,” one noted, “I see no reason not to accept it. But for such a study to be done, you need time: you can’t know in less than one year what the undesirable effects could be.” Another respondent planning to accept vaccination nonetheless expressed uncertainty about safety because of rapid vaccine development: “If I knew that it is effective and safe, but the problem is that I cannot know, because we do not know anything and they have to spend years, they have to experiment with us.”

Qualitative data also showed that those intending to refuse COVID-19 vaccination similarly worried about safety, rapid vaccine development, and side-effects, and their projected refusals were conditional. “I’m too afraid to get a new COVID vaccine,” observed one respondent. “I prefer to wait a bit.” Some 40% of those refusing in France, 18% in Italy and 28% in Spain acknowledged that they might later accept but would “let others get vaccinated first”.

These open responses, too, respondents expressed deep concerns about pharmaceutical companies and about their national governments. “I have no confidence in pharmaceutical laboratories,” one respondent claimed, whereas another contended that these companies have “too many secrets about vaccine ingredients, there’s what’s written, and then what the ingredients really are.”

## Discussion

These results from a large survey in seven European countries show that although up to 66% of respondents anticipate accepting a COVID-19 vaccine, coverage in certain countries and among certain groups may not be sufficiently high to achieve herd immunity [14]. These preliminary results echo other projections of vaccine acceptance in Europe [2, 15-17], and parallel prior theorizations around vaccine confidence, which is closely linked to trust in the vaccine itself, in vaccine producers, and in the health and political structures that promulgate vaccines [8, 9].

Our qualitative and quantitative results indicate that we need to know more than percentages of people reporting vaccine acceptance, and that an understanding of reasons behind vaccine uptake decisions are essential for improving vaccine coverage. The rapid development and specific content of COVID-19 vaccines (e.g. microchips, toxins, adjuvants) and their safety was subject to considerable questioning in our survey, particularly in open text responses, where many respondents conditioned their acceptance and rejection upon evidence of vaccine safety [18]. Such questioning may also reflect citizens’ deeper misgivings about their governments, pharmaceutical companies [7, 19]. Both correlations between projected vaccine acceptance and trust in these authorities and open text responses amply illustrated these misgivings. Moreover, our qualitative evidence emphasized the importance of broad social benefits of accepting COVID-19 vaccination, echoing results elsewhere [15].

Our results have implications for messaging and messengers for COVID-19 vaccination campaigns. Although our survey explored several rumors about COVID-19 and vaccination, it would appear that European governments could achieve greater impact by tackling directly questions about the rapid development and safety of COVID-19 vaccines. Acknowledging public concerns about vaccine contents and the processes by which vaccines were so rapidly developed is essential. Understanding better concerns of more vaccine-skeptical age groups would be important.

Messengers of these explanations are also pertinent to consider. In countries where confidence in the national government competence and honesty is relatively low, governments might find better messengers for vaccine communications than their own officials; working collaboratively with health workers to clarify and explain the safety issues to their patients could be effective, but in some countries where health workers themselves are vaccine hesitant[20], an effort to build bridges with those occupational groups would be essential.

There are two limitations of this preliminary analysis. First, survey panels select representative samples among age and gender groups, regions, and working status, but they draw heavily from those with internet access and of higher educational achievements. Hence, they are not fully representative. In addition, the survey was conducted in December 2020. Since then, the European Medicines Agency has approved other vaccines, and vaccine rollout for specific age groups has begun, although at different rates and with access to different vaccines.

## Conclusion

European public health agencies must tackle COVID vaccine safety concerns head-on so that publics can make informed choices. Communicating clear, proactive messages about scientific advances, safety monitoring and the rapid development of safe vaccines is critical to allaying current concerns. Tailoring these messages to address specific groups and demonstrating the broad social benefits of vaccination can reassure those who are anxious about secondary effects. Finally, given recent reports about health worker refusals for vaccination, European governments might first consider building bridges with health workers, since they are trusted public sources of scientific information. Health workers could be crucial allies in creating and disseminating coherent, coordinated communications in assisting European publics make well-grounded decisions about COVID-19 vaccination.

## Data Availability

Data can be requested from the RECOVER project.

## Acknowledgements

This project is part of the output from RECOVER (Rapid European COVID-19 Emergency research Response), which has received funding from the EU Horizon 2020 research and innovation programme under grant agreement No 101003589.

## Authors’ contributions

LH, MV, BL, NT, and TG-V analyzed the data. TG-V drafted the comment. All authors read, commented on, and added language to the article.

## Data Statement

Data can be requested from the RECOVER project.

